# Text Classification Models for the Automatic Detection of Nonmedical Prescription Medication Use from Social Media

**DOI:** 10.1101/2020.04.13.20064089

**Authors:** Ali Al-Garadi Mohammed, Yuan-Chi Yang, Haitao Cai, Yucheng Ruan, Karen O’Connor, Gonzalez-Hernandez Graciela, Jeanmarie Perrone, Abeed Sarker

## Abstract

Prescription medication (PM) misuse/abuse has emerged as a national crisis in the United States, and social media has been suggested as a potential resource for performing active monitoring. However, automating a social media-based monitoring system is challenging—requiring advanced natural language processing (NLP) and machine learning methods. In this paper, we describe the development and evaluation of automatic text classification models for detecting self-reports of PM abuse from Twitter. We experimented with state-of-the-art bi-directional transformer-based language models, which utilize tweet-level representations that enable transfer learning (e.g., BERT, RoBERTa, XLNet, AlBERT, and DistilBERT), proposed fusion-based approaches, and compared the developed models with several traditional machine learning, including deep learning, approaches. Using a public dataset, we evaluated the performances of the classifiers on their abilities to classify the non-majority “abuse/misuse” class. Our proposed fusion-based model performs significantly better than the best traditional model (F_1_-score [95% CI]: 0.67 [0.64-0.69] vs. 0.45 [0.42-0.48]). We illustrate, via experimentation using differing training set sizes, that the transformer-based models are more stable and require less annotated data compared to the other models. The significant improvements achieved by our best-performing classification model over past approaches makes it suitable for automated continuous monitoring of nonmedical PM use from Twitter.

## Introduction

Prescription medication (PM) misuse/abuse and overdose is a serious, evolving public health problem and a major national health crisis in the United States (US).^1,2^ The Centers for Disease Control and Prevention (CDC) recorded 67,367 drug overdose deaths in the United States in 2018, which resulted from prescription and illicit drug use.^3^ According to the CDC WONDER database records, from 1999 to 2018, more than 232,000 people died in the US from prescription opioids only, with a four-fold increase in that timespan.^4^ Current PM abuse monitoring policies are mostly targeted towards suppliers and licensed doctors, and in most states, patients, prescribers, and drugs distributed are reported for controlled substances through prescription drug monitoring programs (PDMPs).^5^ Law enforcers from the Drug Enforcement Administration (DEA) and prescribers can utilize information from PDMPs to find and restrict possible medication abuse.^6^ However, evidence of the impact of state-level PDMPs is mixed, and current PM monitoring programs are reactive and slow, resulting in a considerable lag between the time a crisis happens and when it is reported.^7–9^ Monitoring mechanisms also lack critical information, such as usage patterns for distinct PMs and user demographics (*e*.*g*., gender and age).^9,10^ Such information can be crucial in designing control measures and outreach programs. Consequently, there is a need for complementary sources of information that can be utilized to develop an effective surveillance systems and protocols.

A number of recent studies have proposed the use of social media for PM and illicit drug abuse monitoring.^11–14^ Data from social media offers a unique opportunity to study human behavior, including behavior associated with the nonmedical use of PMs, at a large scale. It also enables researchers and public health officials to monitor the trends of nonmedical PM use incidents, improve monitoring strategies, and analyze user behaviors.^14–16^ The abovementioned studies have validated that information from social media can be utilized to obtain knowledge about classes of PMs and illicit drugs, typically-used combinations of drugs, etiology of abuse, and populations most affected. The widespread use of social media and the large volume of data that is continuously generated on various social media platforms means that if the relevant information can be efficiently curated, it may be possible to utilize it for obtaining in-depth knowledge about the state of nonmedical PM use and illicit drug use at specific times and places. However, it is not possible to manually curate information from large volumes of data on a continuous basis. Recent advances in computing technology have made it possible to mine very large datasets, such as those available in social media, in close to real time. But the characteristics of text-based health-related information in social media, such as the presence of large amounts of noise and misspellings, and the use of non-standard languages and terms, pose challenges from the perspective of natural language processing (NLP) and machine learning.^17,18^ The task of automatically detecting information about nonmedical PM use, misuse and abuse has been shown to be particularly complex for NLP and machine learning due to factors such as data imbalance (*i*.*e*., only a small portion of the chatter associated with a PM represents self-reports of nonmedical use or abuse), low agreements among manual curators/annotators (*i*.*e*., humans often find it difficult to determine if a user post represents nonmedical use or not), and ambiguous contexts (*i*.*e*., contextual cues indicate nonmedical use, which are detectable by humans but not traditional machine learning models).^9,11^ Consequently, automatic systems, including our past system, for detecting nonmedical PM use from social media have typically shown low performances.^16,19^ Therefore, the development of systems that can automatically detect and filter chatter that represent nonmedical PM use is a fundamental necessity for establishing social media based near real-time monitoring.

In this paper, we model the problem of automatic detection of nonmedical PM use from Twitter data as a supervised classification problem and we present the development of a state-of-the-art classifier that outperforms systems presented in the past. Our proposed classifier is based on context-preserving bidirectional encoder representations from transformers (BERT)^20^— a language representation methodology that has considerably advanced the state-of-the-art in several sentence classification, inter-sentence classification, information extraction (named entity recognition), question answering, and other NLP tasks.^20,21^ When BERT is trained on large unlabeled texts, it is able to capture contextual semantic information in the underlying vector representations, and the representations may then be fine-tuned for several downstream NLP applications. BERT’s key technical improvement is applying the bidirectional training of transformer to language modeling, leading to improved generalizability, enhanced understanding of word meaning, and deep sense of language context and flow. BERT-based models present a significant improvement over past state-of-the-art models that were primarily based on word2vec,^22^ as they represent words or text fragments in a way that captures contextual information causing the same text fragments to have different representations when they appear in different contexts. We also propose fusion learning among multiple BERT-like models to capture additional patterns that may improve classification performance. On a publicly available Twitter dataset with four classes,^23^ our best-performing fusion-based model performs significantly better in terms of detecting PM abuse related posts with an F_1_-score of 0.67 (95% CI: 0.64 – 0.69) than the best traditional classification model, which obtains 0.45 (95% CI: 0.42–0.48). We present an analysis of the system errors to better understand the limitations of BERT-like models, and we recommend future research directions for further improving classification performance on the non-majority PM abuse class. A summary of our contributions is provided below:

i. We propose BERT-based models, fusion learning among multiple BERT-like models, and between BERT-like and deep learning (BiLSTM) models to enhance classification performance for PM abuse detection/classification.
ii. We present extensive performance comparisons of several baseline machine learning, including deep learning, methods with BERT-like models and fusion learning models using a publicly available Twitter PM abuse dataset.
iii. We present empirical analyses of BERT-based models and a discussion of their advantages and drawbacks for application in social media text classification in general and PM abuse detection in particular.

## Materials and methods

### Data collection and annotation

The dataset consisted of tweets mentioning a total of 20 medication-related keywords (*i*.*e*., generic and trade names), which were selected in consultation with a toxicology expert (see Table 1 in supplement). The medications belonged to five classes that are prone to abuse—opioids, benzodiazepines, atypical anti-psychotics, central nervous system stimulants, and gama-aminobutyric acid analogs. In addition to using the drug names as keywords, their commonly occurring misspellings, generated via a data-centric system,^24^ were used to collect tweets via the Twitter public streaming application programming interface (API). The data was annotated by three trained annotators who closely followed a detailed annotation guideline and categorized each tweet into one of four classes(examples of tweets for each category are presented in supplementary information Table 2):

**Table 1.** Performances of traditional machine learning models in terms of class-specific recall, precision and F_1_-scores, and overall accuracy. Best F_1_-score on the *A* class is shown in bold.

| Classification algorithm | Precision | | | | Recall | | | | $F_1$-score | | | | Accuracy (%) |
| --- | --- | --- | --- | --- | --- | --- | --- | --- | --- | --- | --- | --- | --- |
|  | A | C | M | U | A | C | M | U | A | C | M | U |  |
| NB | 0.23 | 0.53 | 0.84 | 0.20 | 0.75 | 0.43 | 0.23 | 0.71 | 0.36 | 0.48 | 0.36 | 0.32 | 38.6 |
| SVM (Setting 1) | 0.60 | 0.70 | 0.74 | 0.80 | 0.31 | 0.62 | 0.89 | 0.74 | 0.41 | 0.66 | 0.81 | 0.77 | 72.3 |
| SVM (Setting 2) | 0.49 | 0.65 | 0.78 | 0.84 | 0.36 | 0.70 | 0.82 | 0.71 | 0.41 | 0.68 | 0.80 | 0.77 | 71.0 |
| RF (n=100) | 0.60 | 0.65 | 0.75 | 0.84 | 0.20 | 0.67 | 0.89 | 0.72 | 0.30 | 0.66 | 0.81 | 0.78 | 71.4 |
| NN [(32,16,8)] | 0.43 | 0.67 | 0.77 | 0.72 | 0.46 | 0.62 | 0.78 | 0.72 | **0.44** | 0.64 | 0.77 | 0.72 | 68.4 |
| KNN (3) | 0.31 | 0.57 | 0.66 | 0.76 | 0.34 | 0.35 | 0.79 | 0.57 | 0.32 | 0.43 | 0.72 | 0.65 | 58.8 |

**Table 2.** Performances of deep learning models in terms of class-specific recall, precision and F_1_-scores, and overall accuracy. Best F_1_-score on the A class is shown in bold.

| Classification algorithm | Precision | | | | Recall | | | | $F_1$-score | | | | Accuracy (%) |
| --- | --- | --- | --- | --- | --- | --- | --- | --- | --- | --- | --- | --- | --- |
|  | A | C | M | U | A | C | M | U | A | C | M | U |  |
| CNN (Twitter-GloVe-embedding) | 0.47 | 0.65 | 0.75 | 0.76 | 0.39 | 0.61 | 0.82 | 0.69 | 0.43 | 0.63 | 0.79 | 0.72 | 69.03 |
| Char-CNN | 0.36 | 0.63 | 0.72 | 0.57 | 0.32 | 0.50 | 0.81 | 0.58 | 0.34 | 0.56 | 0.76 | 0.58 | 64.04 |
| **BiLSTM**<br>**Twitter-GloVe-**<br>**embedding** | 0.44 | 0.70 | 0.77 | 0.76 | 0.45 | 0.60 | 0.82 | 0.76 | **0.45** | 0.65 | 0.80 | 0.76 | 70.01 |

1. *Potential abuse or misuse* (*A*): The tweet has potential evidence that the user is nonmedically using, misusing or abusing the medication, or is expressing the intent to.
2. *Non-abuse consumption* (*C*): The tweet specifies that the user has a valid prescription for the medication and is taking the medication as prescribed or is seeking to obtain the medication for a valid indicated reason.
3. *Drug mention only* (*M*): The tweet mentions the medication but there is no evidence of consumption.
4. *Unrelated* (*U*): The tweet mentions a medication-related keyword but is referring to something else.

A total of 16,443 tweets were annotated into the above classes. The distribution of each class is as follows: A=2,636 tweets, C=4,589, M=8,563, and U=655. The tweets were divided into 11,829 training, 1,343 validation, and 3,271 held-out test data as shown in Figure 1. Further details regarding the preparation and availability of the data is available with our past publication.^23^

**Figure 1:**
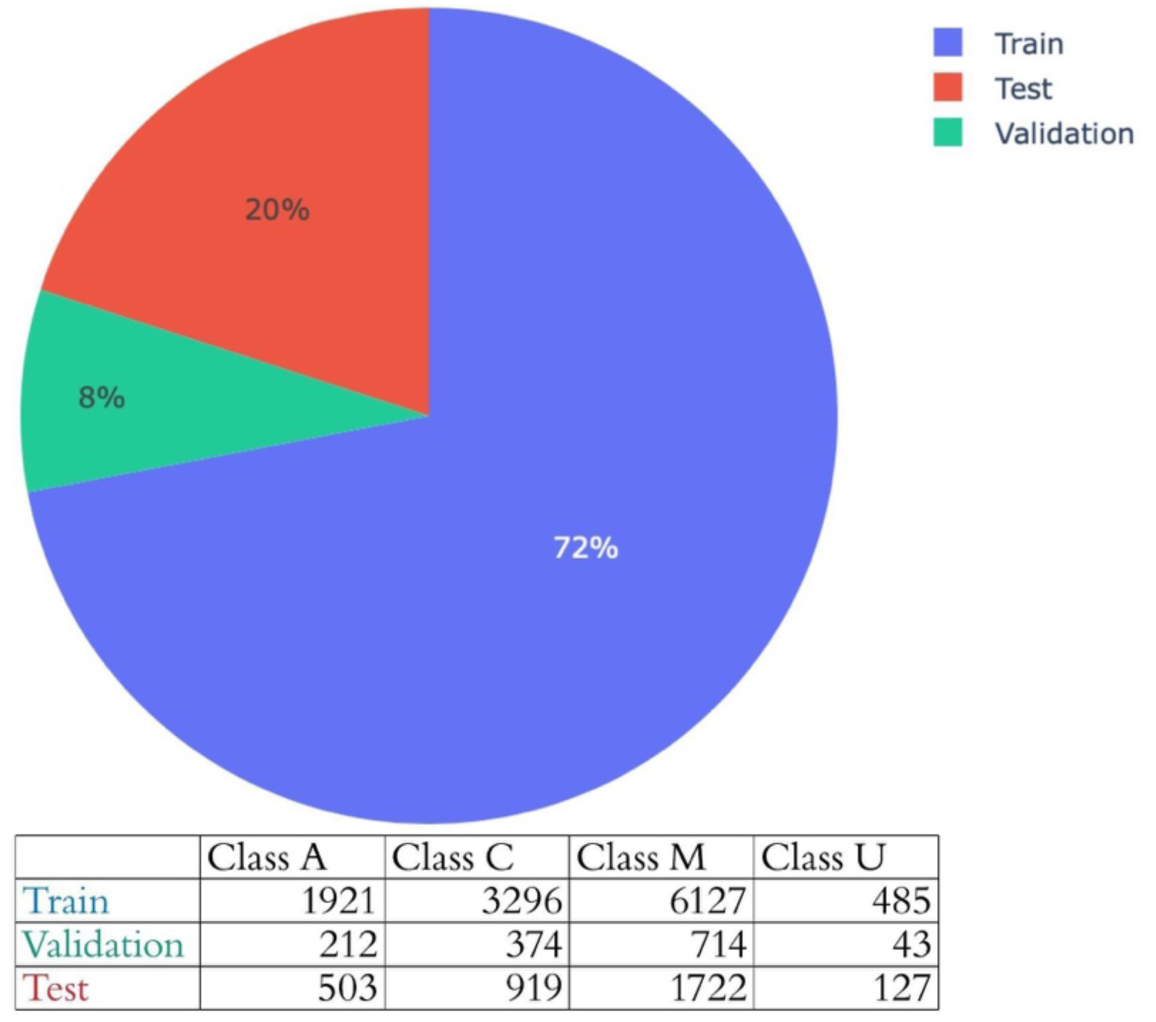
Class distribution in the training, validation, and test datasets.

### Experiment design and implementation

We experimented with multiple classifiers and compared their performances on the test set. We categorized the classifiers into three broad sets. All classifiers were trained on the same training set and hyper parameters were optimized using the same validation set. We describe them in the following subsections.

#### Traditional machine learning models

The first set of classifiers, which we refer to as *traditional*, consisted of non-deep neural network classifiers from distinct families, as described by Fernandez-Delgado et al,^25^ to mitigate selection bias and obtain a comprehensive baseline. These included support vector machines (SVM), random forests (RF; decision tree family), Gaussian naïve bayes (NB; Bayesian family), shallow neural networks (NN; neural network family), and k-nearest neighbor (nearest-neighbors family), We used the above classifiers as baselines for the classification with the following configurations:

- **SVM** (setting1: with radial basis function as kernel)^26,27^
- **SVM** (setting2: linear SVM trained using stochastic gradient descent)^28^
- **RF** (100 trees)^29^
- **Gaussian NB**^30^
- **Shallow NN** (three hidden layers), and
- **k-nearest neighbor (KNN)** (restricted to three neighbors)^31^

The tweets were first pre-processed by lowercasing, removing URLs, user names, and non-English characters, stemmed by the Porter stemmer, and then converted to features: n-grams (contiguous sequences of words with n ranging from 1 to 3), and word clusters (a generalized representations of words learnt from medication-related chatter collected on Twitter),^32^ each with the most frequent 1,000 items.

#### Deep learning models

For several years, deep learning methods have achieved state-of-the-art results in many NLP topics, including multiclass text classification. For deep learning approaches, we used the three commonly used deep learning approaches for text classification as follows (all models hyper parameters are chosen based on their optimal performance on validation dataset, these hyper parameters are presented in supplementary information Table 3):

**Table 3.** Performances of transformer-and fusion-based models in terms of class-specific recall, precision and F_1_-scores, and overall accuracy. Best scores for each metric over all the classifiers shown in bold

| Classification algorithm | Precision | | | | Recall | | | | $F_1$-score | | | | Accuracy (%) |
| --- | --- | --- | --- | --- | --- | --- | --- | --- | --- | --- | --- | --- | --- |
|  | A | C | M | U | A | C | M | U | A | C | M | U |  |
| **BERT_base** | 0.60 | 0.78 | 0.86 | 0.88 | 0.61 | 0.77 | 0.85 | 0.89 | 0.60 | 0.77 | 0.86 | 0.89 | 79.48 |
| **BERT_large** | 0.60 | 0.79 | 0.86 | **0.91** | 0.61 | 0.77 | 0.86 | 0.85 | 0.61 | 0.78 | 0.86 | 0.88 | 79.85 |
| **RoBERTa_large** | 0.63 | 0.81 | 0.88 | 0.90 | 0.66 | 0.82 | 0.87 | 0.89 | 0.65 | 0.81 | 0.88 | **0.90** | 82.32 |
| **AlBERT_xxlarge** | 0.66 | 0.81 | 0.88 | 0.86 | 0.63 | 0.83 | 0.88 | 0.88 | 0.65 | 0.82 | 0.88 | 0.87 | 82.78 |
| **XLNet** | 0.65 | 0.77 | 0.86 | 0.87 | 0.55 | 0.83 | 0.86 | 0.82 | 0.60 | 0.80 | 0.86 | 0.85 | 80.52 |
| **DistilBERT** | 0.56 | 0.75 | 0.86 | 0.89 | 0.60 | 0.77 | 0.83 | 0.87 | 0.58 | 0.76 | 0.84 | 0.88 | 78.0 |
| **Proposed Fusion-1** | 0.60 | **0.84** | **0.91** | 0.78 | **0.76** | 0.81 | 0.84 | **0.93** | **0.67** | 0.82 | 0.87 | 0.85 | 82.22 |
| **Proposed Fusion-2** | 0.67 | 0.83 | 0.87 | 0.88 | 0.62 | **0.83** | **0.90** | 0.89 | 0.65 | **0.83** | **0.89** | 0.88 | 83.43 |
| **Proposed Fusion-3** | 0.56 | 0.83 | 0.90 | 0.75 | 0.73 | 0.80 | 0.83 | 0.92 | 0.64 | 0.82 | 0.86 | 0.82 | 80.92 |
| **Proposed Fusion-4** | **0.68** | **0.84** | 0.87 | 0.89 | 0.62 | 0.82 | **0.90** | 0.87 | 0.64 | **0.83** | **0.89** | 0.88 | **83.49** |

- **Convolutional neural networks (CNN)**^33^: A CNN is a deep learning architecture that is frequently used for hierarchical document classification.^34^
- **Character-level CNN (Char-CNN)**^35^: Char-CNN for text classification is effective for text classification and robust to misspelling or word variation that cannot be captured by word level CNN or BiLSTM.
- **BiLSTM with Twitter GloVe word embeddings**^36^: One of the dominant NN architectures for text classification is the recurrent neural network (RNN)^37,38^ Most cutting-edge RNN architecture for text classification use the BiLSTM architecture,^39^ and the first input layer is word embeddings. Given the word-level data, we used Twitter GloVe word embeddings,^36^ which is generated from 2B tweets and 27B tokens, contains 1.2M vocabulary, is uncased, and has 200d vectors.

#### Transformer-based models

Variants of transformer based models have very recently emerged as the state-of-the-art in many NLP tasks,^20,40–42^ but their performance for complex health-related social media datasets, such as our dataset for PM abuse, have not been thoroughly evaluated. In such models, the transformer structure removes the recurrent connections that are present in BiLSTM architectures, and with such an architecture, transformer-based models such as BERT can be trained on very large amounts of data efficiently, and then be fine-tuned for downstream NLP applications. The success of BERT-like models arises from the fact that they can capture bi-directional contextual information (*i*.*e*., the vector representations of words, or character sequences, depend on preceding and following words). Thus, unlike GloVe and word2vec, BERT embeddings preserve contexts, thus, enabling the vector-based representation of long sequences of characters or sentences. Figure 2 illustrates how this property helpful for our target task as the word ‘*drugs*’ has a different embedding representation based on the context of each sentence/tweet.

**Figure 2.**
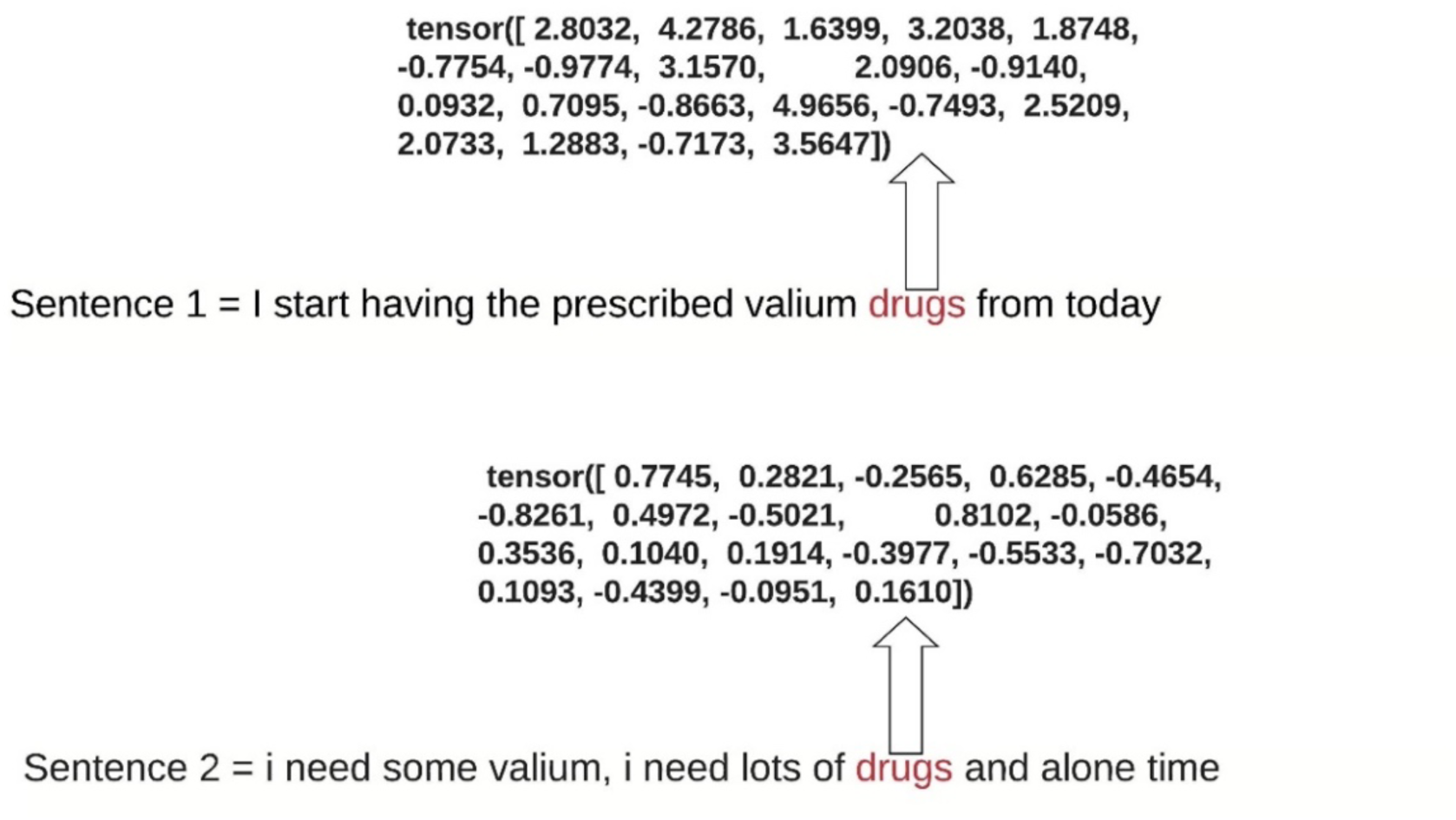
Sample sequence embeddings showing how the term “drugs” is represented differently when appearing in different sequences.

In our experiments, we used several BERT variants and fusion-based approaches that combine, for each sentence/tweet, the outputs from multiple models to predict the class. All model hyper parameters are chosen based on their optimal performance on validation dataset, these hyper parameters are presented in supplementary information Table 3. The models are as follows:

- ***BERT-1***^20^: We used the original BERT base, which consists of 12 layers (transformer blocks), 768 hidden size 12 attention heads with total of 110M parameters.
- ***BERT-2***^20^: We used original BERT-large, which consists of 16 layers (transformer blocks), 1024 hidden size 16 attention heads with total of 340M parameters.
- ***RoBERTa***^40^: RoBERTa is a variant that employs an enhanced process for training BERT models, overcoming problems with undertraining. It has outperformed BERT in several NLP tasks.
- ***AlBERT***^41^: This is a light version of BERT that has achieved new state-of-the-art results on several NLP benchmarks with fewer parameters compared with BERT-large.
- ***XLNet***^42^: The authors in 35 proposed XLNet, a generalized autoregressive pretraining method that overcomes the BERT limitations using autoregressive approach. They showed that XLNet outperforms BERT on several NLP tasks.
- ***DistilBERT***^43^: This is a small, general-purpose language model that can be fine-tuned for specific NLP tasks and has shown performances comparable to larger BERT-based models.
- ***Proposed Fusion-1***: Fusing probabilities of each tweet from BERT-2, AlBERT, and RoBERTa (base classifiers) using a NB classifier (metaclassifier).
- ***Proposed Fusion-2***: Fusing probabilities of each tweet from BERT-2, AlBERT, and RoBERTa (base classifiers) using a logistic regression classifier (metaclassifier).
- ***Proposed Fusion-3:*** Fusing probabilities of each tweet from BiLSTM, AlBERT, and RoBERTa (base classifiers) using a NB classifier (metaclassifier).
- ***Proposed Fusion-4:*** Fusing probabilities of each tweet from BiLSTM, AlBERT, and RoBERTa (base classifiers) using a logistic regression classifier (metaclassifier).

### Evaluation

Since our overarching objective is to develop a system for detecting self-reports of PM abuse from streaming Twitter data, our primary metric for comparing classifiers was the F_1_-score (harmonic mean of precision and recall) for the *A* (abuse/misuse/nonmedical use) class. Precision, recall and F_1_-score are computed as shown in below equations (1, 2, and 3). To determine statistical significance in performance differences, we computed the 95% confidence intervals (CIs) for the F_1_-scores using the bootstrap resampling technique with 1000 resamples.^44^ In addition, we also computed recall, precision, F_1_-scores for the other classes to investigate if any notable differences in performances across the four classes could be observed, and the overall accuracies of the classifiers (% of correct predictions).

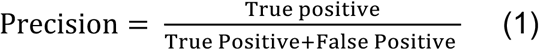

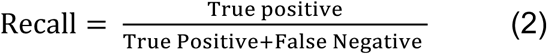

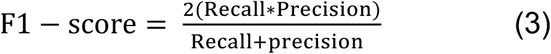

## Results

### Traditional machine learning models

Table 1 presents the results of the traditional machine learning classifiers on the held-out test set. The NN classifier yields the highest F_1_-score for A (0.44; 95% CI: 0.41-0.47). As indicated by 95% CI range, this classifier is significantly better at detecting PM abuse-related posts compared to the NB, RF, and kNN classifiers, but not the SVM classifiers. The SVM classifier with a radial basis function kernel (setting 1) obtained the highest accuracy (72.3%), although the table shows that this accuracy is driven by superior performances over the other classes. Interestingly, the best F_1_-score obtained for the A class is comparable to the one we observed in our prior work (0.46), where we modeled the problem as a binary classification task and used a much smaller training data.^16^

#### Deep learning models

Table 2 presents the results of deep learning-based classifiers on the held-out test set. The RNN with BiLSTM attention mechanism and Twitter GloVe embeddings obtained the best F_1_-score of 0.45 (95% CI: 0.42-0.48). Its performance is comparable to the performance of the CNN classifier with GloVe embeddings, but significantly better than the char-CNN classifier. Similar performance patterns can be observed for the accuracies as well. Despite the significantly more computational power needed by the deep learning classifiers compared to the classifiers presented in Table 1, there are no significant improvements in performances. This finding is also consistent with our past research on similar problems.^13^

#### Transformers and fusion based models

The results presented in Table 3 illustrate strengths of BERT-based models for this classification task compared to all the classifiers presented in Tables 1 and 2. Fusing the probabilities obtained from multiple classifiers led to consistently improved F_1_-scores ranging between 0.67 to 0.64 compared to single transformer-based approaches whose scores ranged from 0.58 to 0.65. The best F_1_-score (0.67; 95% CI: 0.64-0.69) was obtained by the proposed fusion-1 approach, which was significantly better than all the methods from Tables 1 and 2. The fusion-based approaches also performed better than or at least as good as the single transformer-based models for all other metrics and all the classes, although the differences were not always statistically significant.

## Discussion and Post-classification Analyses

Our experiments verified the significant differences between transformer-based models and previous state-of-the-art machine learning, including deep learning, models. The low proportion of abuse/misuse representing tweets caused past classifiers to perform poorly on this class, obtaining F_1_-scores similar to the ones presented in Tables 1 and 2 and Figure 3. While some studies reported good overall accuracies or micro-/macro-averaged F-scores over all the classes, these metrics are primarily driven by the majority class(es), and do not reflect the performances of the approaches on the important non-majority class. From our experiments, it is evident that (i) transformer-based approaches considerably improve PM abuse detection (in our case, by approximately 20 points in terms of F_1_-score as shown in Figure 3); and (ii) a fusion-based approach that combines the outputs of multiple transformer-based models is more robust in terms of performance than a single transformer-based method. The second finding is unsurprising since past research involving traditional classifiers have shown that ensemble learning typically outperforms individual classifiers^45^. The improved classification performance, however, comes with a heavy cost in computing time, particularly during training, as the hyper-parameters of each model needs to be fine-tuned to optimize performance.

**Figure 3:**
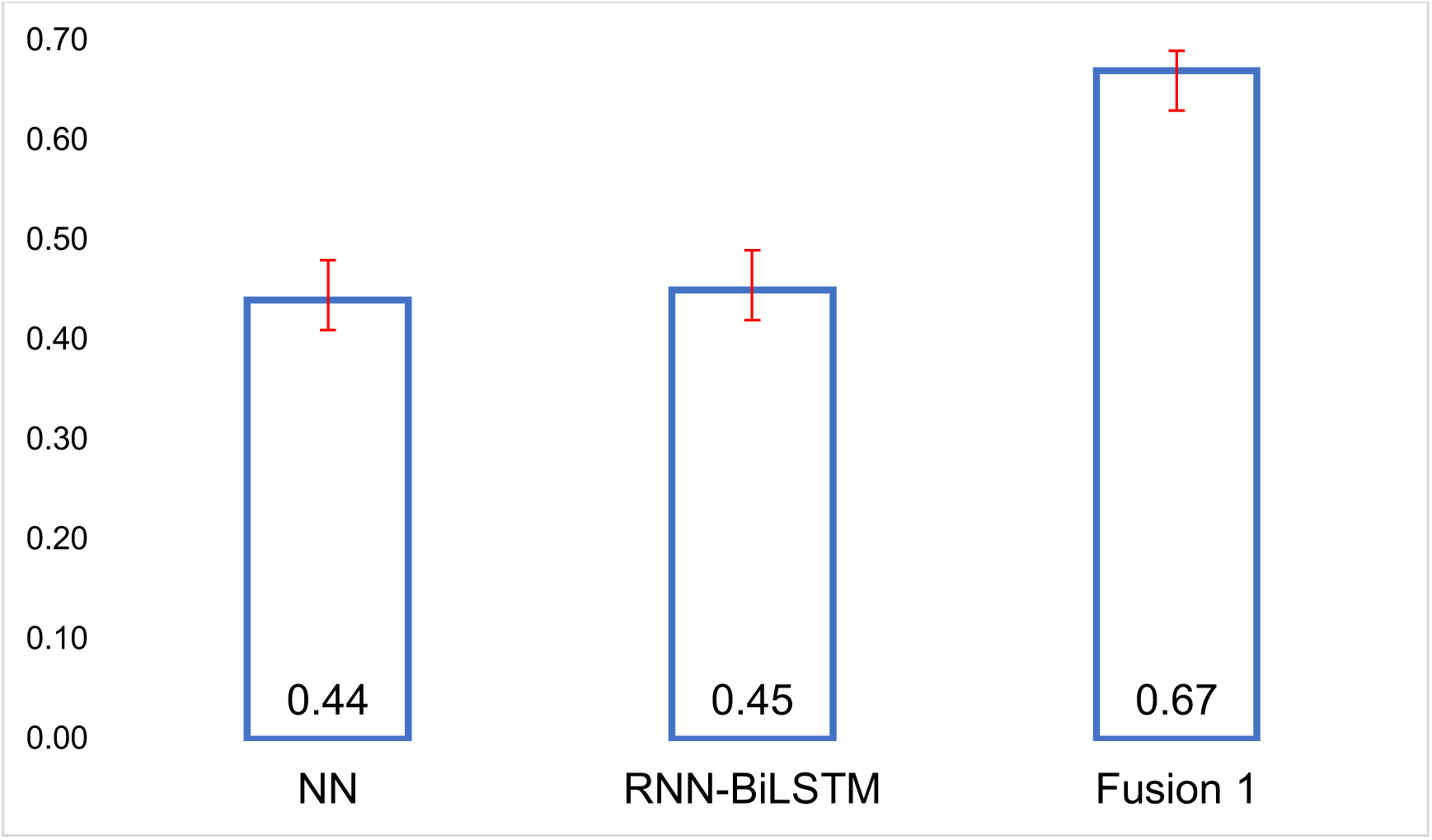
Comparison between the best models from each learning category (A F1-score Class A)

### Analysis of the effect of training size on model performance

We repeated the classification experiments by varying the size of the training set and evaluating each classifier on the same test set. Our intent was to determine how the performances of the classifiers varied with training set sizes, particularly the rates of increases in performances with increases in training set sizes. We drew stratified samples of the training set consisting of 25%, 50%, and 75% of all the tweets, and computed the F_1_-scores over class A for all the classifiers.

Figure 4 shows the performances of the classifiers for the different training set sizes. The figure reveals two key information: (i) even with very small training data set sizes, transformer-based models consistently outperform other models, and (ii) while some of the traditional machine learning models appear to hit a ceiling in performance, transformer-based models appear to keep improving as more training data is added. Finding (i) illustrates the strength of such models, particularly in terms of text representations, which enable relatively high performances even with small amounts of training data. Finding (ii) is promising as it suggests further improvements to classification performances are possible.

**Figure 4:**
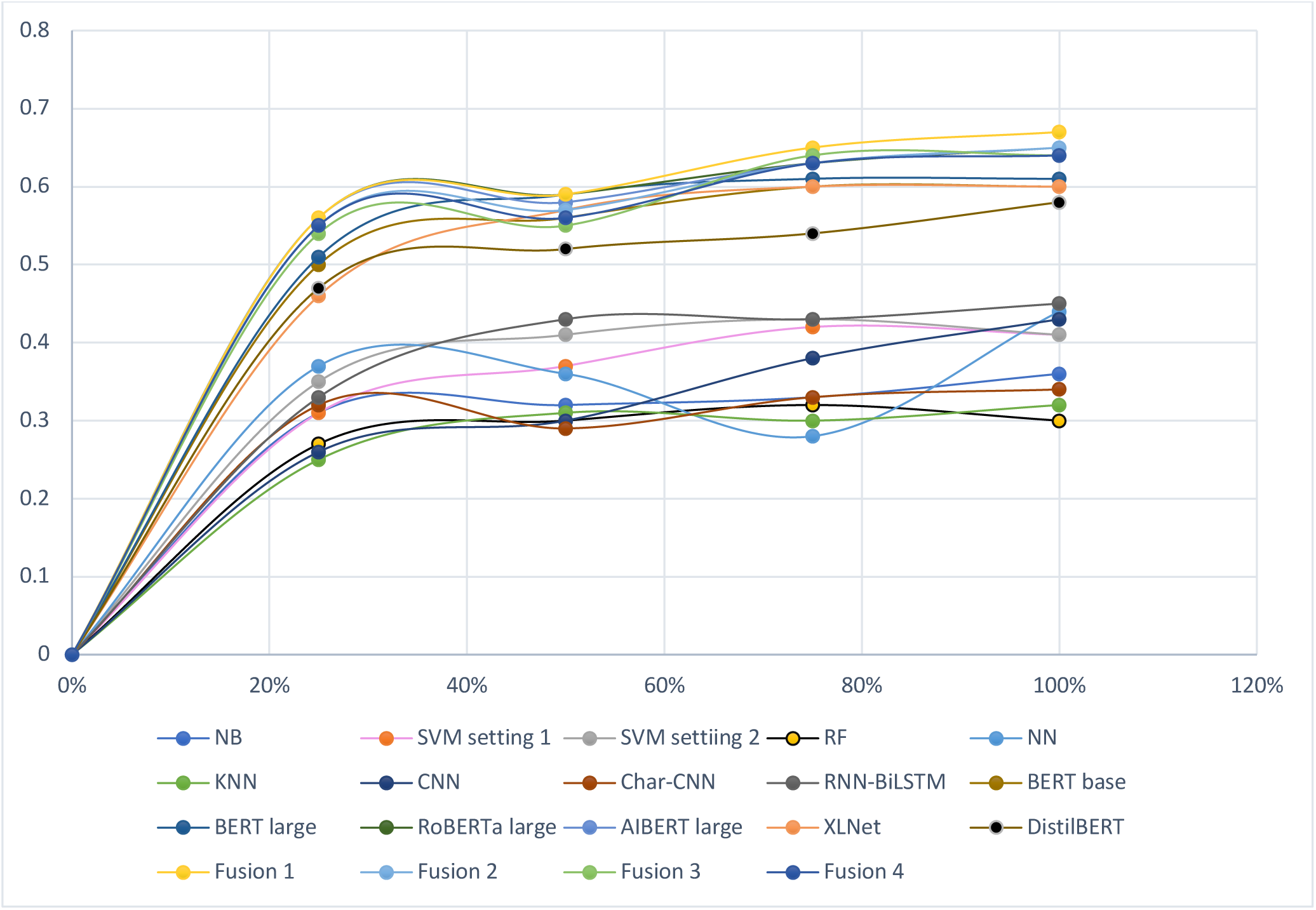
Learning curve at different Amount of Training Data Used for Training.

#### Post classification content analysis

We classified 100k tweets using the developed model, then we performed content analysis by calculating the term frequency–inverse document frequency (TFIDF) as shown in Figure 5. The high TFIDF terms give us an overview of the contents in each class. Our objective was to assess and verify if the classification strategy actually managed to separate tweets representing different contents. For example, in the abuse chatters (class A), we see that the content involves tweets are reporting how the user abuse PM, such as usage of more than typical dosage (*taking mixing*) or PM co-consumption (*Whiskey*), and the reason for misuse, such as for recreation (*first-time, took-shit*). The high-frequency topics in PM consumption chatters (class C) indicate the users using the medications for medical symptoms (*panic-attacks, mental health, nerve pain*), or discussing side effects they experienced (*side-effect*). Interestingly, the dominant theme in PM mention chatters (class m) is related to opioid crisis (*drug-legal, prescription-needed, purdue-pharma*), and a variety of other topics regarding the medications (*powerful-prescription, college-grades, need-powerful*) that require further analysis to understand the specific information. This surface-level topic analysis suggests that the classifier is indeed able to distinguish contents associated with abuse or nonmedical use of the PMs.

**Figure 5:**
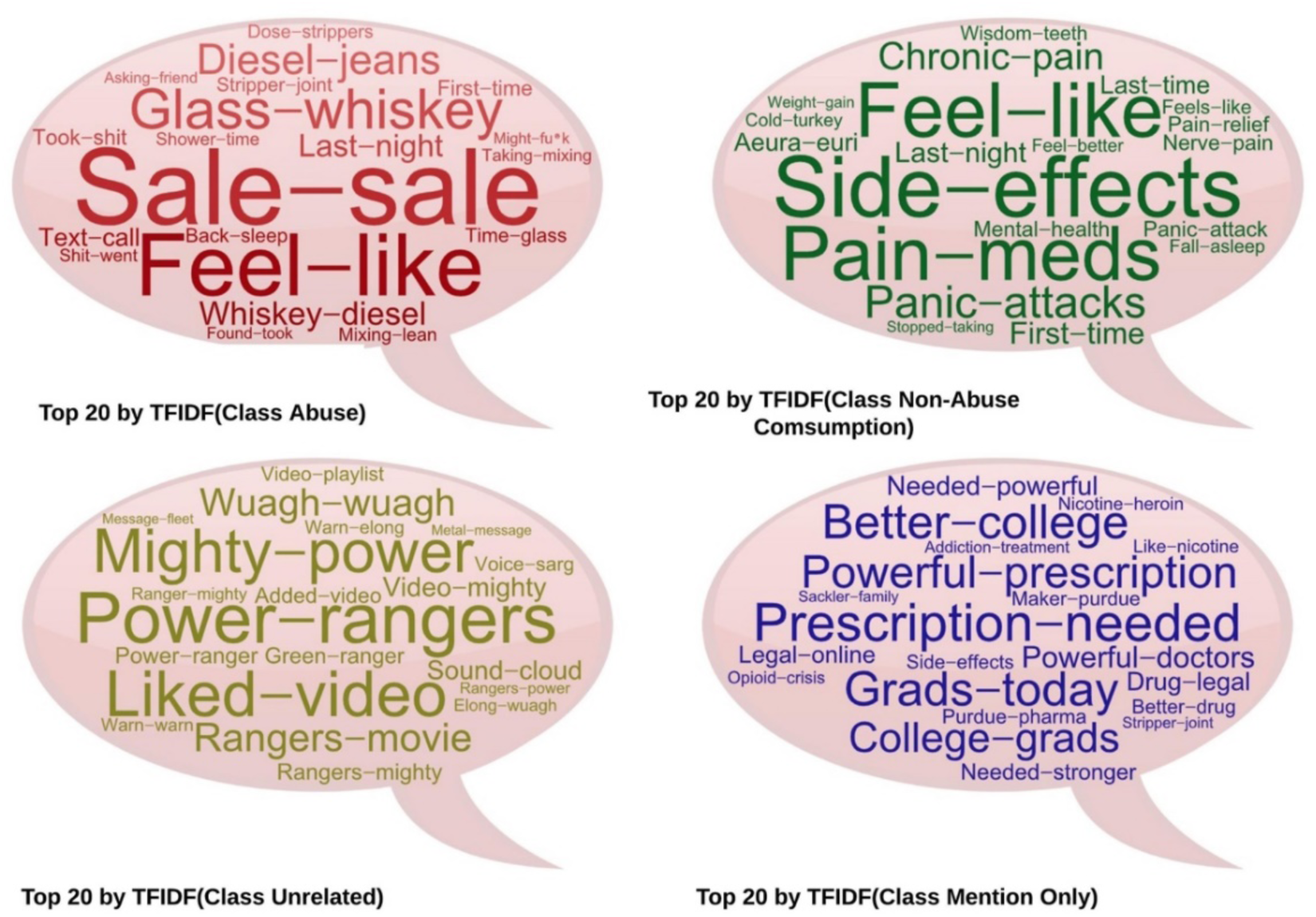
20 TF-IDF bigram word clouds for automatically classified tweets from each of the 4 categories.

### Challenges and possibilities for improving classification performance

Building on our promising experimental findings, we analyzed the errors made by our best-performing classifier in order to identify the common causes of errors (as shown in the confusion matrix in Figure 6) and to explore possible mechanisms by which performance may be further improved in future research.

**Figure 6.**
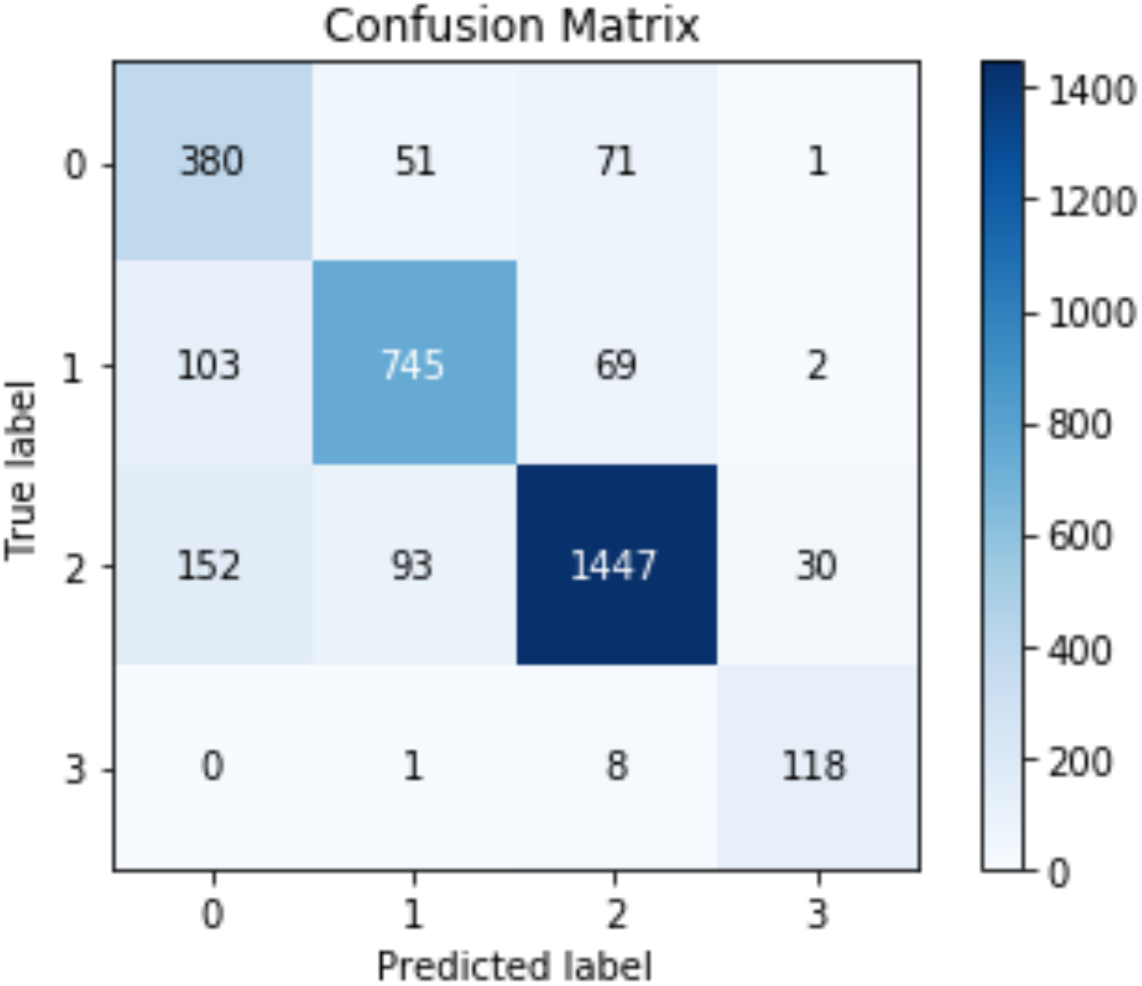
Confusion Matrix Fusion 1 based Model (0=Class A, 1=Class C, 2= Class M, 3=Class U)

#### Lack of complete context

While BERT-like models are able to capture context-dependent meanings of words better, they still lack the *commonsense/pragmatic inference*^46^ capabilities of humans. When it comes to PM abuse/misuse detection, the mechanism of action of a medication is purely human knowledge and is not necessarily explicitly encoded in tweets. In the example below, the human annotator recognizes that Adderall® is used for performance enhancement by students, and hence can infer that the tweet represents *possible* misuse/abuse. However, due to the lack of explicit contextual cues, the machine learning classifier mis-classifies the tweet as *M* (*i*.*e*., medication mention).

*Example 1: @userX1 books, @userX2 Adderall, @userX3 the place*.

While there is no existing method for capturing commonsense, it may be possible to add further context to a given tweet by incorporating additional tweets by the same user (*e*.*g*., tweets posted before and after the given tweet).

#### Absence of deep connections within the context

Humans can generalize more and observe connections deeper than those presented in the context or said words. Humans connect the word within said sentences with speakers’ social groups, culture, and age, as well as their life experience to obtain full understanding, which might be even difficult for humans from different backgrounds, ages, or social groups to understand. In examples 2 and 3, while the annotators were capable of connecting the relationship between Adderall® and laundry, and a movie (*Harry Potter* in this case) and Xanax®, our classifiers failed to classify them correctly.

*Example 2: laundry and adderall go side by side*

*Example 3: we see harry potter and pop Xanax*

Such tweets represent extremely difficult cases for the classifiers, particularly if there are no other similar tweets annotated in the training set. In addition to annotating more data, which is always a time-consuming yet effective method for improving machine learning performance, future research may try to incorporate more user-level information (*e*.*g*., details from the user’s profile, followers, following *etc*.) to improve performance.

#### Influence of the pretraining dataset

The BERT model used in this study was pre-trained using a corpus of books (800M words) and English Wikipedia (2,500M words),^20^ and the other models were pre-trained on similar datasets. However, social media language differs from that in books or Wikipedia, and in past research using word2vec, we have observed that word embedding models trained specifically on social media data improved system performances for social media text classification tasks.^47^ Therefore, conducting pre-training using large social media datasets may help improve performance further.

#### Users’ network influence on understanding the context

Beyond the issue of language context, social media network features, such as mentioning or retweeting, can affect the meaning of sentences. Consequently, capturing the meaning and the context only from the text is challenging. For example, writing content inside quotation marks implies that the statement is not the user’s and is quoted from another user:

*Example 4:* “*someone send me xanax i’ll pay”*

*Example 5: someone mail me xanax i’ll pay*

Both examples were classified by all BERT-based models as abuse. However, example 4 was considered nonabuse by human annotators because it was represented between two quotations and just mentioned what somebody else says. Example 5 was considered abuse and represented the user himself/herself. Misclassification possibly occurred because the quoted statements were observed much less in the training dataset compared with general statement. In a scenario where the patterns are evident but rarely represented in the training dataset, incorporating a simple rule that can guide the algorithm in understanding this situation or similar situations can improve performance, although the example is not well represented in the training.

#### Related work

Recent efforts for the analysis of social media text for studying PM and drug abuse can be categorized into three groups on the basis of the methodology employed: (i) manual analysis; (ii) unsupervised analysis, and (iii) automatic classification using supervised machine learning. In most early works, researchers proposed and tested hypotheses via manual analyses of social media contents (e.g., examining opioid chatter from Twitter to determine the presence of self-reports of misuse/abuse). Chan et al.^48^ used two weeks’ of data from Twitter to manually code users’ messages and message contexts (personal versus general experiences). Similarly, Shutler et al ^49^ aimed to qualitatively examine tweets that mentioned prescription opioids to decide if they represent abuse or non-abuse, whether they were characterizable, and to examine the connotation (positive [i.e., analgesic use], negative [i.e., adverse event], or non-characterizable). The second approach is unsupervised methods, which have been popular for finding trends from large social media datasets, such as applying topic modeling using LDA ^50^ to identify topics that are associated with selected drugs. However, past researcher^51^ demonstrated that only small amounts of data may present abuse information, and the unsupervised methods are probably considerably affected by unrelated content. Consequently, the decisions derived might be unreliable or generalized. When working with general social media data, developing and applying a robust supervised classification approach before topic modeling or trend analysis could improve the conclusion derived from this approach to understand the text and is methodologically more robust.^51^

The third approach is supervised machine learning, particularly automatic text classification, and it enables researchers to overcome the problems associated with unsupervised methods by filtering out unrelated content. However, supervised machine learning methods need high-quality, manually annotated datasets to train, and, if the trained models show promising results, they can be applied to large datasets to curate relevant data automatically. Multiple distinct approaches have been attempted for automatically detecting drug abuse/misuse from social media chatter. For example, Jenhani et al.^52^ developed hybrid linguistic rules and a machine learning-based approach to detect drug-abuse-related tweets automatically. In our past work,^16^ we aimed to investigate the opportunity of using social media as a resource for the automatic monitoring of prescription drug abuse by developing an automatic classification system that can classify possible abuse versus no-abuse posts. In some studies,^53 54^, deep learning models were developed to detect drug abuse risk behavior using two datasets. The first dataset was manually annotated and a deep learning model trained on the first dataset was applied to annotate the second dataset automatically. Both datasets were then used to train and develop the final deep learning model. Some studies have used social media sources other than Twitter,^55^ employing machine learning methods (LR, SVM, and RF) to determine whether a Reddit post was about opioid use disorder recovery. Despite the potential application of supervised classification approaches, our recent review on the topic^51^ showed that significant improvements in the performances of current systems were needed to effectively utilize social media data for PM abuse monitoring.

## Conclusion

Developing an effective PM abuse detection system for social media data holds substantial practical application in establishing a drug abuse surveillance system that can complement traditional mechanisms of monitoring. A social media-based system will also enable close to real-time analyses, and, perhaps, the early detection of potential future crises like the current opioid crisis. In this study, we built on state-of-the-art NLP methods, particularly transformer-based pre-trained models such as BERT, to significantly improve automatic detection of PM abuse from Twitter over past approaches. We ran extensive experiments and compared the performances of multiple machine learning methods, including deep learning methods, with BERT-like models and fusion learning models using a large annotated Twitter PM abuse classification dataset. We also show that by employing a fusion-based classifier that combines prediction from multiple models, the classification performances can be further improved and made more stable. Our analyses of the system performances and misclassifications revealed possible future research tasks that may further improve performance.

## Data Availability

Training data has been made public. Details in the following link:
https://sarkerlab.org/pm_abuse_data/

## Author contributions statement

MAA, YCY, HC, YR and AS conducted the data collection, analysis, classification and evaluations. MAA conducted most of the classification training and evaluation. JP provided medical domain expertise and supervision for the study. GG and JP provided high-level guidance/supervision for the study. AS designed the annotation objectives and the overall study. MAA, YCY, GG, JP and AS contributed to the manuscript writing.

## Acknowledgments

Research reported in this publication is supported by the NIDA of the NIH under award number R01DA046619. The content is solely the responsibility of the authors and does not necessarily represent the official views of the NIH.

